# Estimation of the Ascertainment Bias in Covid Case Detection During the Omicron Wave

**DOI:** 10.1101/2022.04.22.22274198

**Authors:** Ahmed Elbanna

## Abstract

Covid cases in the general population have been historically underreported due to a variety of reasons including limited access to PCR testing at the start of the pandemic, lack of nation-wide surveillance testing, and discouraged testing unless symptomatic. Concerns about underreporting have increased during the Omicron surge due to the expanded use of at-home rapid tests which are not required to be officially reported. For the state of Illinois, we have found that reported cases constituted only 50%-70% of the actual cases during the pre-Omicron waves (August 2020-December 2021). During the first Omicron (BA1) wave, this fraction dropped to 20-29% (i.e., only 1 in 4 to 1 in 5 cases are reported). During the ongoing second Omicron (BA2) surge, this fraction has further decreased to 12-18% (i.e., only 1 in 6 to 1 in 8 cases are reported). These estimates have important implications on understanding the extent of the Omicron surge at the state and national levels.

## Methodology

We compare incidence rate in faculty/staff at UIUC where extensive surveillance data is available and the detected case rate is close to the actual one, to the case rate at the state level where there may be significant underreporting. We use this connection to estimate the underreporting at the state levels in all the epidemic waves given some knowledge of the actual case rates at the state and campus levels at only one point during Fall 2020. Specifically, we proceed as follows:

## Step 1: Estimating case detection ratio at the state level in Fall 2020

The best estimate for ascertainment bias on the state level comes from data in August/September 2020 when access to testing had improved compared to the early days of the pandemic and vaccines were not deployed yet. To estimate the underreporting in cases during that period, we looked at two important figures:

1. Case fatality Ratio (CFR): This is defined as the ratio of 7-day average daily deaths at time t to the 7-day average of confirmed cases at time t-21 days.

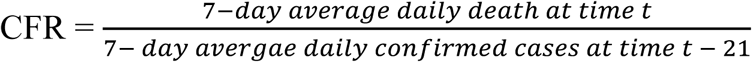

Note that both the numerator and denominator in this ratio are known sufficiently well. Using the Johns Hopkins database (through divoc-91 website)^1^ the CFR in August/September 2020 was estimated to be on average equal to 1%.

2. Infection fatality Ratio (IFR): This is defined as the ratio of 7-day average daily deaths at time t to the 7-day average daily actual number of cases at time t-21 days.

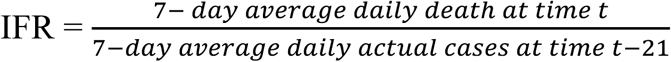

The first estimate for infection fatality ratio (IFR) came from Wuhan^2^ in January 2020 to be on average 1%. As more data was collected from around the world, it became clear that the infection fatality ratio has a steep age dependence^3^. By late summer 2020, the estimate for the population-wide averaged infection fatality ratio (IFR) most probably ranged between 0.5% and 0.7%^3^.

With these two numbers we compute the ascertainment ratio in case detection during that period as follows:

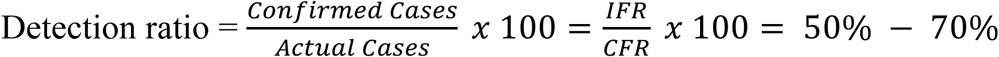

## Step 2: Comparing confirmed case rate for faculty/staff to case rate at the state level

Our next step is to relate cases on campus, where surveillance testing is available, to cases in the state. Data on cases for faculty/staff at UIUC is accessible through the campus dashboard^4^. Cases at the state level are recorded by IDPH^5^. We have found a strong correlation between cases in the faculty/Staff cohort and cases in the state. Figure 1 illustrates this remarkable correlation. Interestingly, the case rate per 100k in faculty/staff agrees reasonably well with the confirmed case rate per 100k for the general population in the state for all the pre-Omicron waves.

**Figure 1:**
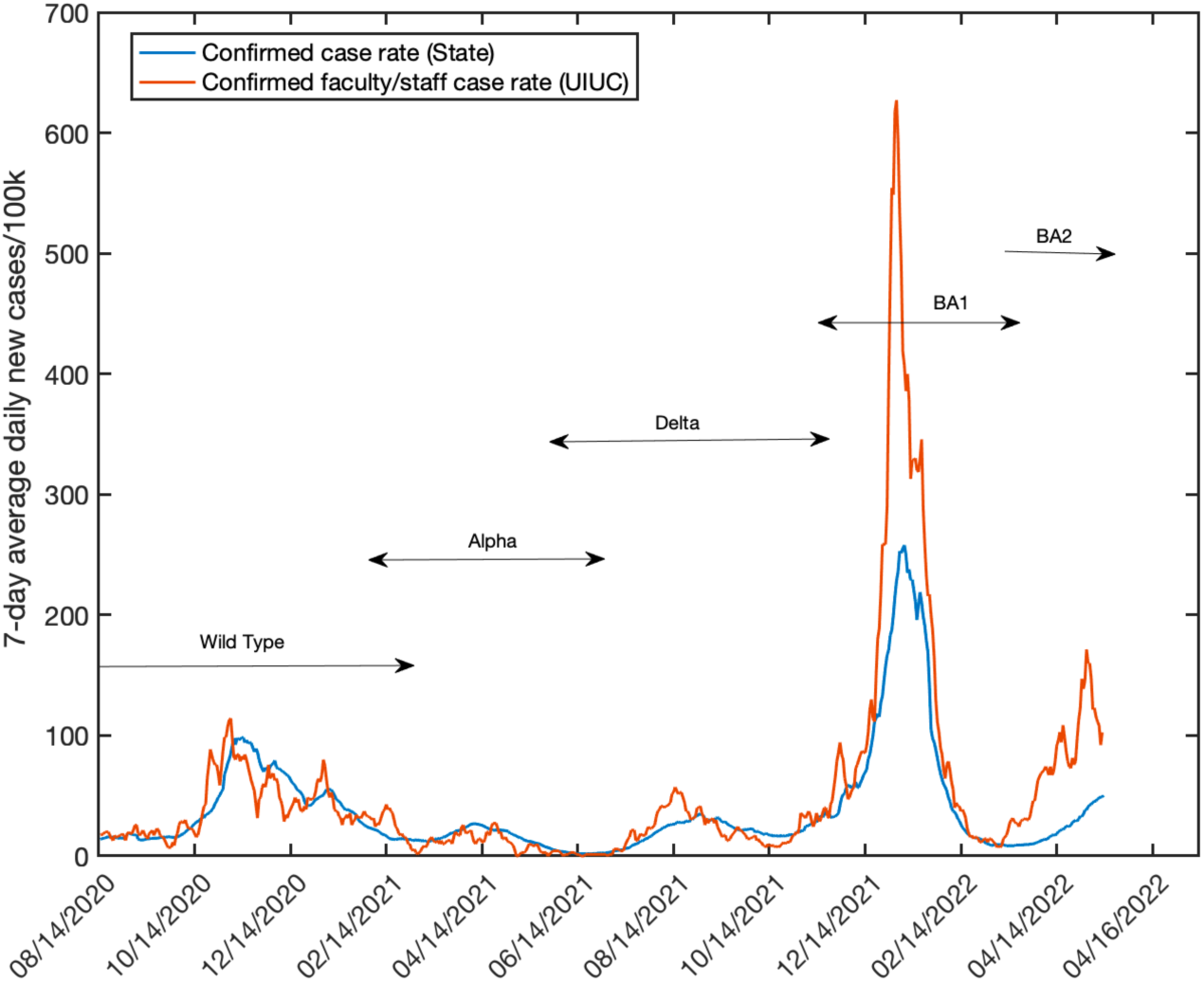
Confirmed case rate per 100k for faculty/staff at UIUC compared to the confirmed case rate per 100k at the state level. Actual case rate at the state level is higher due to the higher ascertainment bias (see text for details)

We assert that the detected case rate in the faculty/staff cohort is very close to their actual case rate. This is because during 2020-2021 academic year, UIUC was testing everyone who needed campus access. In Fall 2021, while mandatory testing was no longer required for vaccinated faculty/staff, the convenient covidShield test was still widely accessible and most faculty/staff took advantage of this. Therefore, while it is possible that the detected case rate in faculty/staff during the Delta and Omicron waves is different from the actual case rate, we may still assume that this difference is small and that the detected case rate may be taken as a proxy for prevalence in this subpopulation. This assertion gives us an opportunity to determine the ratio between faculty/staff case rate and the case rate at the state level.

## Step 3: Establishing the relation between actual case rates at the state and campus levels

By using the detection ratio above (for Fall 2020) and the observation that the actual case rate in faculty/staff is equal to the confirmed case rate at the state level during that period, we arrive at the following conclusion

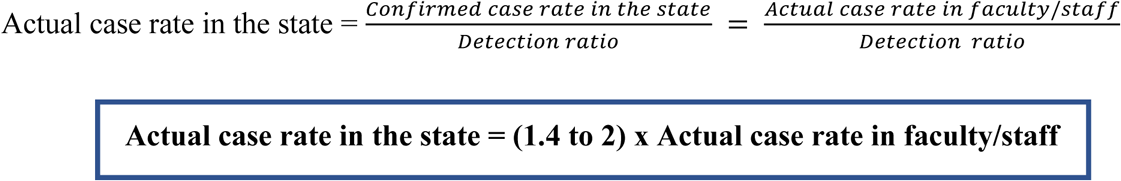

This is the key relation that we will be using in estimating the underreporting at the state level by tracking case rate in faculty/staff. This relation should approximately hold at any time during the pandemic. This is because the exposure patterns and the social dynamics of faculty/staff are similar to those of the general population. What may change, however, is the fraction ***f*** of actual cases at the state level that gets officially reported.

## Step 4: Estimating the fraction f of actual cases at the state level that is officially reported

We are now at a position to estimate this fraction ***f*** for the different epidemic waves.

### 1. Pre-Omicron waves (August 2020-December 2021)

Confirmed case rate in the state = Actual case rate in faculty/staff

Actual case rate in the state = (1.4 to 2) x Actual case rate in faculty/Staff

= (1.4 to 2) x Confirmed case rate in the state

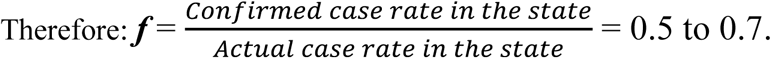

That is, approximately, only 1 in 2 cases (or perhaps only 2 in 3 cases) was reported at the state level before the Omicron surge.

### 2 Omicron BA1 wave (Dec 2021-January 2022)

Confirmed case rate in the state = 0.41 x Actual case rate in faculty/staff

Actual case rate in the state = (1.4 to 2) x Actual case rate in faculty/staff

= (3.4 to 4.9) x Confirmed case rate in the state

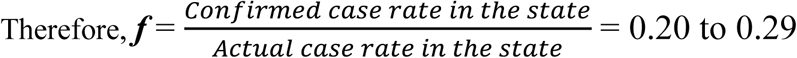

That is, approximately, only 1 in 4 or 1 in 5 cases was reported at the state level during the first Omicron surge.

### 3 Omicron BA2 wave (April 2022)

Confirmed case rate in the state = 0.26 x Actual case rate in faculty/staff

Actual case rate in the state. = (1.4 to 2) x Actual case rate in faculty/staff

= (5.4 to 7.7) x Confirmed case rate in the state

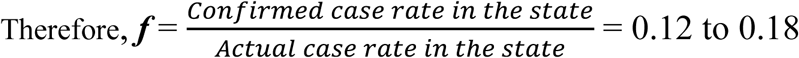

That is, approximately, only 1 in 6 to 1 in 8 cases is reported at the state level during the ongoing Omicron BA2 surge. This reduction in reporting is consistent with the expanded use of rapid tests, the limited access to PCR testing due to closure of some free testing sites, as well as the role of vaccination or prior infection in reducing the symptoms to a milder degree thus discouraging seeking PCR testing.

## Conclusions

1-During pre-Omicron waves (August 2020-December 2021): 1 in 2 cases got to be reported officially at the state level

1. -During Omicron BA1 wave (December 2021-January 2022): Only 1 in 4 or 1 in 5 cases got to be reported officially at the state level.
2. -During Omicron BA2 wave (April 2022): Only 1 in 6 or 1 in 8 cases are reported officially at the state level. This is also consistent with the CDC surveillance data^6^ that suggests the use of rapid at home tests were tripled between late 2021 and March 2022
3. -Since 08/14/2020, the State of Illinois has reported a total of 3.024 million Covid19 confirmed cases. The actual number of people who got infected during the same period is estimated to range between 6.424 and 8.576 million cases. That is, on average 35%-47% of the cases were officially reported. However, the underreporting varied widely with time. Figure 2 shows the variation in the confirmed cases rate per 100k at the state level as a function of time compared to the estimated actual case rate.

**Figure 2:**
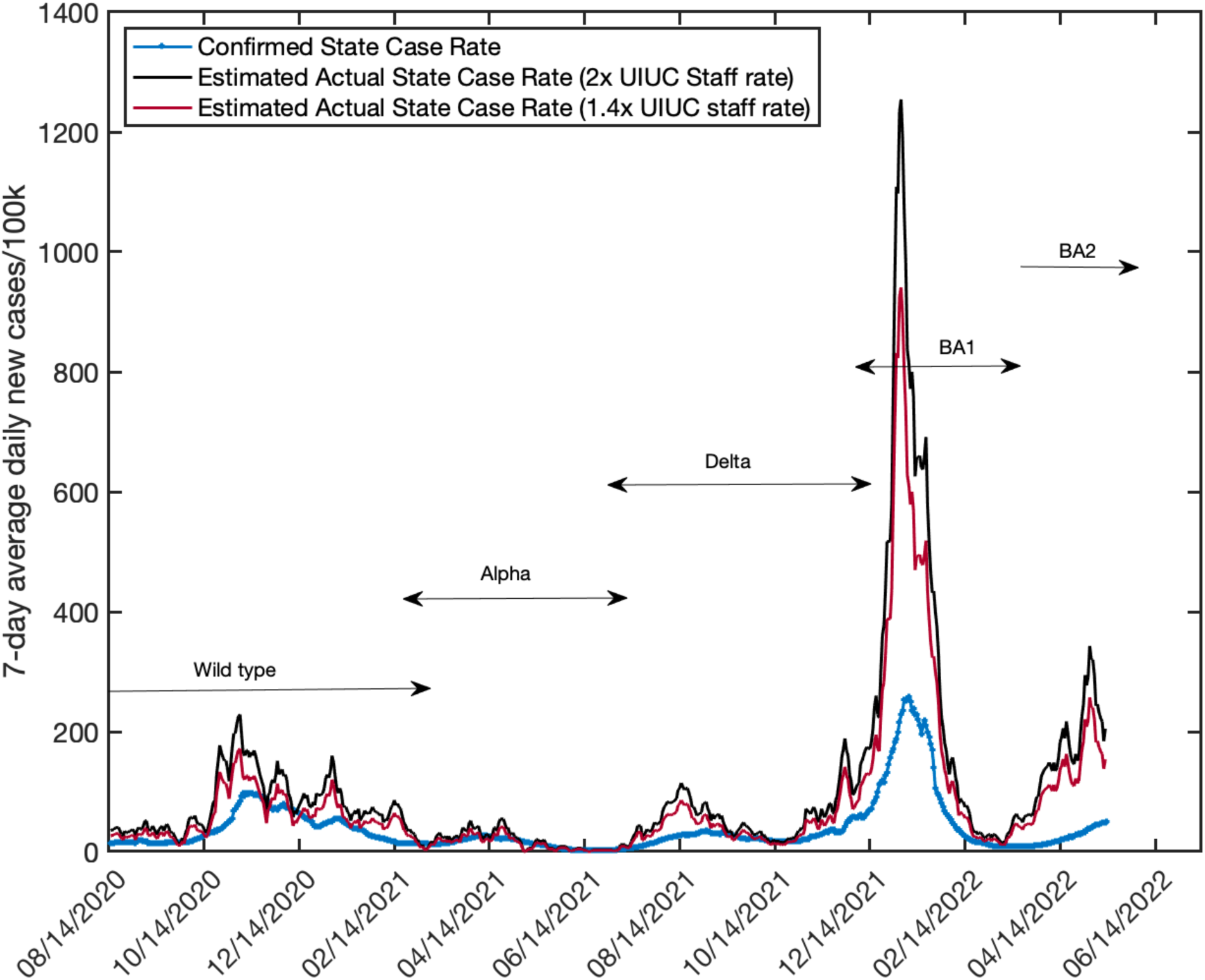
Confirmed case rate per 100k at the state level compared to the estimated actual case rate that ranges between 1.4x-2x the case rate in faculty/staff at UIUC. During the peak of Omicron BA1 wave, only 20-25% of the cases were officially reported. During the current Omicron BA2 wave, only 12-18% of the cases are officially reported.

## Data Availability

All data used is available online at
UIUC COVID-19 dashboard archived database: https://uofi.app.box.com/s/nrrnx2fn6sqt4hhuh2ckq2u6z5o4zmqg/file/878498867995
IDPH COVID-19 data on daily cases, tests and deaths: https://dph.illinois.gov/covid19/data/data-portal/cases-tests-and-deaths-day.html

## Acknowledgement

A.E acknowledges the UIUC Shield testing program which conducted more than 2.7 million tests on the Urbana-Champaign campus since July 2020 and continues to provide high-quality surveillance data on COVID-19 transmission dynamics.

